# COVID-19 seropositivity changes in asymptomatic individuals during the second and third waves of COVID-19 in Tokyo

**DOI:** 10.1101/2020.09.21.20198796

**Authors:** Sawako Hibino, Kazutaka Hayashida, Andrew C Ahn, Yasutaka Hayashida

## Abstract

**Importance:** The decline of anti-SARS-COV-2 antibody titers has been documented. It is not clear whether the antibodies are persistent in infected individuals who had no symptoms. Serial serological tests on the same individuals can provide insights into the population-level prevalence and dynamic patterns of seropositivity against COVID-19 infection.

**Objective:** To assess changes in COVID-19 seroprevalence and the persistence among the asymptomatic working population in Tokyo from May 2020 to December 2020, spanning from the second wave to the third wave of COVID19 through serial testing on the same individuals.

**Design:** We conducted a cohort observational study about SARS-COV-2 seropositivity on the same individuals with no COVID-19 symptoms from May 26 to December 15, 2020. Six hundred fifteen healthy volunteers (mean ± SD 40.8 ± 10.0, range 19 - 69; 45.7 % female) from 1877 office workers were initially enrolled from 11 disparate locations across Tokyo. Participants having fever, cough, or shortness of breath at the time of testing were excluded. Fingertip blood was applied onto the cassette of a COVID19 IgM/IgG rapid test kit to detect antibodies. We compared two test results with the same kits approximately one month apart for the self-reference to validate the positive test results. Initial two tests were performed weekly from May 26 to August 25. Those who had positive test results either at the first or second test were offered the follow-up test on Dec 8 or 15, 2020. An outside ethical committee reviewed and approved the protocol.

**Participants:** Healthy office workers from 11 disparate locations (1877 employees in total) across Tokyo volunteered to the study. Participants having fever, cough, or shortness of breath at the time of testing were excluded.

Main Outcome(s) and Measure(s): Seropositivity rate (SPR) was calculated by pooled data from each two-week window from May 26 to August 25. Either IgM or IgG positivity was defined as seropositive. Changes in immunological status against SARS-CoV-2 were determined by comparing results between two tests a month apart obtained from the same individual. A Follow-up test was offered to the seropositive individuals on December 8 or 15.

**Results:** Six hundred fifteen healthy volunteers (mean + SD 40.8 + 10.0; range 19 - 69; 45.7 % female) received at least one test. Seropositivity rate (SPR) increased from 5.8 % to 46.8 % during the second wave. The most dramatic increase in SPR occurred in late June and early July, paralleling the rise in daily confirmed cases within Tokyo, which peaked on August 4. Out of the 350 individuals (mean + SD 42.5 + 10.0; range 19 - 69; 46.0 % female) who completed the initial two tests, 152 participants were found to be seropositive at either the first or second test. Out of 152 seropositive individuals, 74 participants (52%: women, median age: 44 years, range: 23 – 69 years) underwent the third test. The interval between the initial positive results and the third test was approximately four months (mean ± S.D. 120 ± 17 days). Thirty participants (40.5 %) became seronegative at the third test on December 8 or 15.

**Conclusions and Relevance:** COVID-19 infection may have spread widely across the general population of Tokyo despite the quite low fatality rate during the second wave. Given the temporal correlation between the rise in seropositivity and peaking in reported COVID-19 cases that occurred without a shut-down, Tokyo might achieve herd immunity temporally at the second wave. Substantial reduction of the seropositivity among asymptomatic individuals in four months in December may explain why Tokyo had the third wave, the resurgence of COVID19 is occurring every 3 to 4 months, and the herd immunity strategy has not succeeded.

**Summary:**

- Serial testing of antibodies in the same individuals with a month apart provided self-reference to validate the results for asymptomatic individuals.
- The weekly seropositivity rate among the participants increased from 5.8 % (before) to 46.8 % (during) in parallel to the increase of confirmed case number of COVID-19 during the second wave of COVID-19 in Tokyo.
- Participants who had positive results in the tests a month apart had follow-up test four months after the positive results.
- Among the seropositive participants, 40.5 % turned seronegative at the follow-up test.
- The temporal correlation between the rise in the seropositivity and the decrease in reported COVID-19 cases without imposing lockdown measures during the second wave may indicate Tokyo temporarily achieved herd immunity.
- Substantial reduction of seroprevalence in the asymptomatic population in four months may explain why the third wave hit Tokyo, the resurgence of COVID19 is occurring every 3 to 4 months, and the herd immunity strategy has not succeeded.

## Introduction

Mortality from COVID-19 has been low in Japan as compared to the United States and European countries^1^. The reasons for the low number of deaths are unknown. They may relate to either the low prevalence of SARS-CoV-2 infections in the general population or diminished fatality rates among those infected. Distinguishing which of these factors comes into play requires data on the total prevalence of COVID-19, particularly among the asymptomatic general population. To estimate the incidence of COVID-19, serology tests were obtained in asymptomatic individuals throughout the summer of the year 2020, incorporating the time before, during, and after the well-documented “second-wave” of COVID-19 infection in Japan. For each participant, serology tests were offered twice a month apart to provide self-references of test results and to estimate the seroprevalence of the general population in Tokyo during the second wave. Furthermore, the decline of anti-SARS-COV-2 antibody titers has been documented^2^. It is unclear whether the antibodies are persistent in infected individuals, especially those who had no symptoms. The follow-up serological tests on the same individuals can provide insights into how the seropositivity among the asymptomatic population changes over time.

## Methods

Six hundred fifteen healthy volunteers (mean ± SD 40.8 ± 10.0, range 19 - 69; 45.7 % female) out of 1877 employees of a large Japanese company from 11 disparate locations across Tokyo were enrolled. As is the general practice in Japan, participants commuted daily to their workplace: remote working was not common. First, each participant was offered two tests about one month apart. These tests were performed weekly from May 26 to August 25 (except for 6/2 and 8/11; those designated to 8/11 were rescheduled to 8/18). Participants having fever, cough, or shortness of breath at the time of testing were excluded. Fingertip blood was applied onto the cassette of the COVID19 IgM/IgG rapid test kit (Aurora Biomed, Vancouver, Canada) to detect antibodies. The seropositive case numbers were combined every two weeks, and Seropositivity Rates (SPR) were calculated. The SPR 95 % confident interval (95% C.I.) was calculated by binomial distribution [±1.96×√ (p(1-p)/n)]. We compared the first two test results a month apart for the self-reference to validate the results. Further, those who had positive test results were offered the follow-up test on Dec 8 or 15. An outside ethical committee reviewed and approved the protocol.

### Patient and Public Involvement

Patients who had previously contracted COVID-19 were not involved in setting the research question, design, or outcome measures.

## Results

The demographic characteristics of the participants were composited every two weeks and summarized in Table 1. Seroprevalence increased from 5.8 % to 46.8 % throughout the summer (Figure). The most dramatic increase in SPR occurred in late June and early July, paralleling the rise in daily confirmed cases within Tokyo^3^, which peaked on August 4. Out of the 615 participants, 350 individuals (mean ± SD 42.5 ± 10.0; range 19 – 69; 46.0 % female) completed the initial two tests. The interval between these tests was 30.5 ± 5.6 days (Mean ± S.D.). Among 350 individuals, 152 had seropositive results. 93.2 % (142/152) of these seropositive individuals showed some status change between the two tests (e.g., seronegative to positive, or seroconversion) (Table2). 12.2 % (12/98) of the seropositive participants at the first test became seronegative at the second test (Table 2). There were no instances where the two tests a month apart revealed physiologically unexpected changes – for example, a case where IgM negativity and IgG positivity became IgM positive in a month. Among 152 seropositive participants, 74 participants (52%: women, median age: 44 years, range: 23 – 69 years) underwent the follow-up test. The interval between the test showing positive results and the third test was approximately four months (mean ± S.D. 120 ± 17 days). Thirty of 74 participants (41 %) were seronegative at the third test (Table 3).

**Figure.**
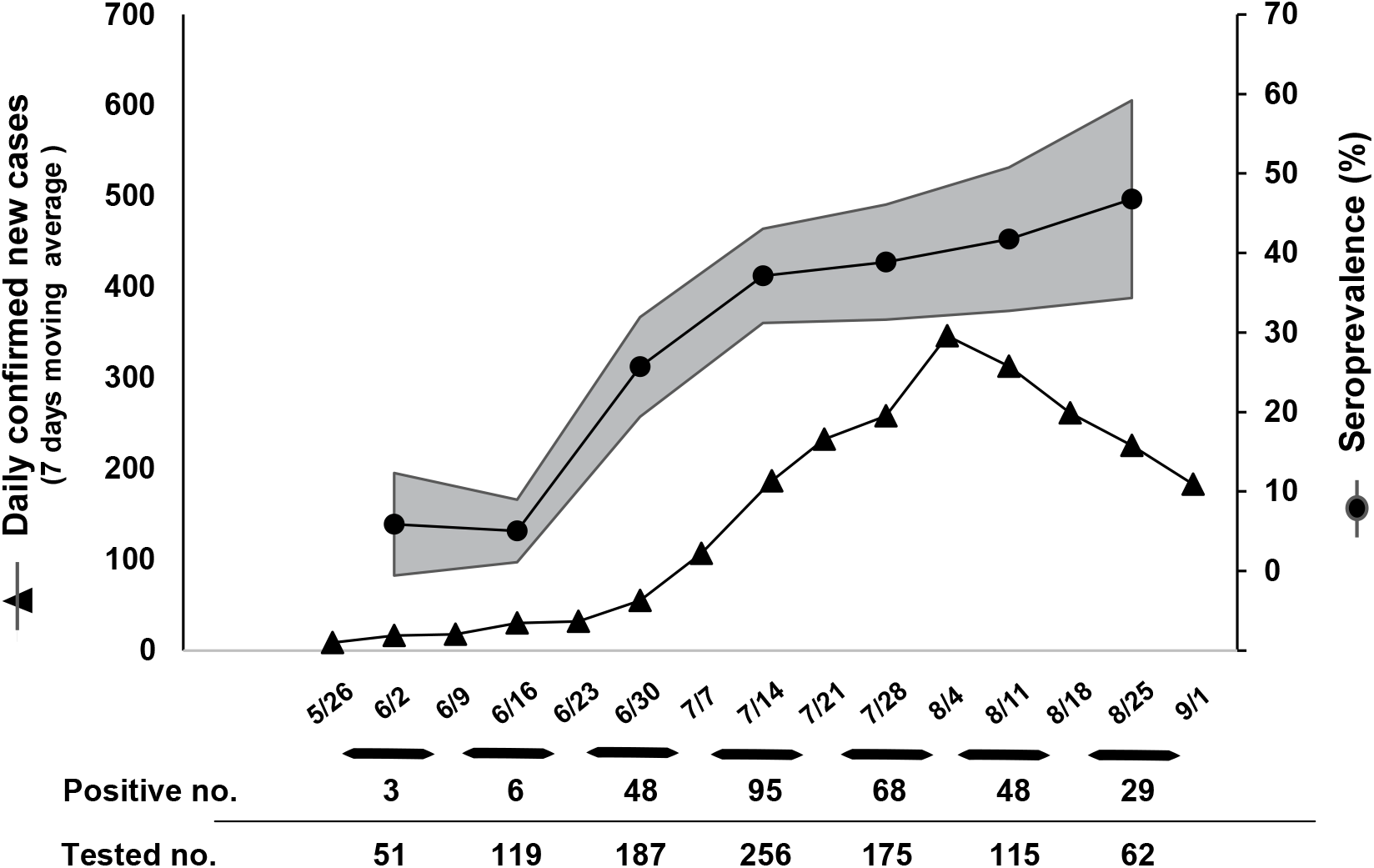
The seropositivity rate and 7 days moving average of daily confirmed new cases of COVID-19 during the second wave. Tests were performed at 5/26, 6/9, 6/16, 6/23, 6/30, 7/7, 7/14, 7/21,7/28, 8/4, 8/18 and 8/25. Data from every two weeks are combined, and SPR is calculated. SPR (closed circle) with 95 % confident interval (95% C.I.) is plotted on the graph. 7 days moving average of daily confirmed new case number in Tokyo on the indicated dates (closed triangle) is also shown. Seropositivity numbers and the total number of antibody tests performed for each two-weeks window are shown at the bottom of the graph.

**Table 1.** Baseline background characteristic of the participants on each two-weeks window.

| Date | 5/26-6/9 | 6/10-6/23 | 6/24-7/7 | 7/8-7/21 | 7/22-8/4 | 8/5-8/18 | 8/19-9/2 |
| --- | --- | --- | --- | --- | --- | --- | --- |
| Age , mean (SD) | 45.6 (9.1) | 42.5 (8.7) | 40.7 (9.5) | 40.9 (10.3) | 41.0 (10.5) | 41.5 (9.7) | 41.6 (12.2) |
| No. (%) |  |  |  |  |  |  |  |
| Age groups, y |  |  |  |  |  |  |  |
| 30> | 3 (5.9) | 9 (7.6) | 27 (14.4) | 39 (15.2) | 28 (16.0) | 10 (8.7) | 12 (19.4) |
| 30-39 | 9 (17.6) | 36 (30.3) | 59 (31.6) | 79 (30.9) | 50 (28.6) | 43 (37.4) | 18 (29.0) |
| 40-49 | 23 (45.1) | 50 (42.0) | 70 (37.4) | 85 (33.2) | 62 (35.4) | 40 (34.8) | 15 (24.2) |
| 50-59 | 13 (25.5) | 20 (16.8) | 26 (13.9) | 44 (17.2) | 28 (16.0) | 18 (15.7) | 13 (21.0) |
| 60 or older | 3 (5.9) | 4 (3.4) | 5 (2.7) | 9 (3.5) | 7 (4.0) | 4 (3.5) | 4 (6.5) |
| Gender |  |  |  |  |  |  |  |
| Male | 43 (84.3) | 74 (62.1) | 104 (55.6) | 137 (53.5) | 82 (46.9) | 59 (51.3) | 23 (37.1) |
| Female | 8 (15.7) | 45 (37.8) | 83 (44.4) | 119 (46.5) | 93 (53.1) | 56 (48.7) | 39 (52.9) |

**Table 2.**
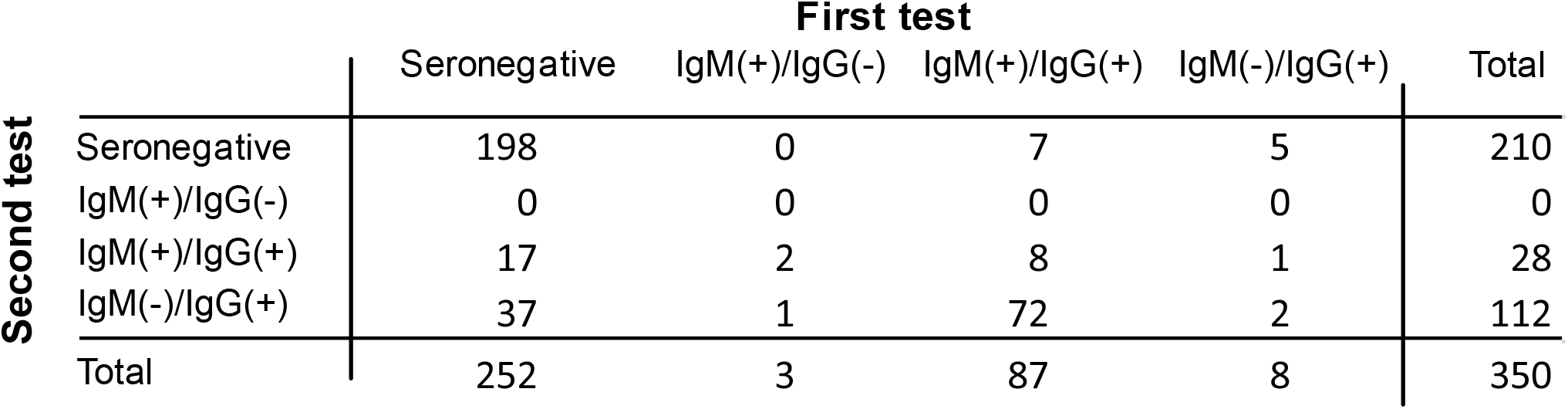
The comparison of two test results a month apart in the same individuals.

**Table 3.**
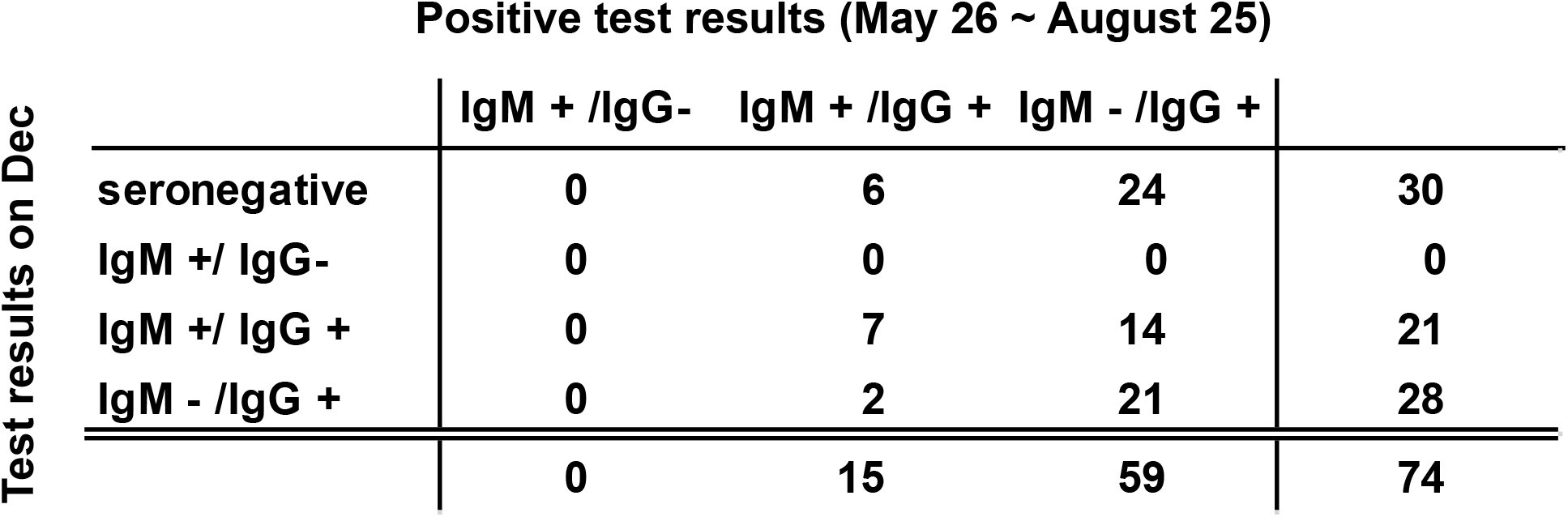
The comparison of first antibody positive results and test results on Dec in the same individuals.

## Discussion

Seroprevalences against COVI-19 in Japanese populations have been reported, which shows a wide variety of results (0.03%-3.3%)4,5, as we see similar considerable variations among seroprevalence studies against COVID-19 in general6. These variations probably arise from the difference in thresholds among individual test kits6. The challenge to determine the exact COVID-19 seroprevalence in the asymptomatic population is that we are unsure whether the same threshold for acute symptomatic cases can be applied to asymptomatic patients. We used the first generation of rapid antibody test kit (older version) from Aurora Biomed, which uses a mixture of COVID-19 S and N antigen for antibody capture. It probably helps to increase the detectability and may have a lower threshold. However, this also usually increases the false-positive result rate, although the company claimed 100 % specificity in their document.

Therefore, we validate the test results by comparing the two test results a month apart for self-references. The comparison confirmed the rapid kit did not yield any unreasonable answers: there were no illogical transitions (e.g., IgM-/IgG+ to IgM+/IgG-), almost all seropositive individuals showed antibodies status change in a month. Thus, we concluded that the positive results are valid, although the observed SPR as high as 46.8 % in our cohort is exceptionally high compared to the seroprevalence of COVID-19 in a known epicenter like New York City (20.2 %)7. Soon after our cohort SPR reached 40 %, the second wave peaked without imposing strict “lockdown” measures. These observations may indicate that Tokyo had a temporary herd immunity at that time.

We performed follow-up tests on these seropositive individuals on December 8 and 15, right after the third wave of COVID-19 started. Only half of the seropositive participants (74/152) underwent the third test. However, more than 40 % of them turned seronegative in four months. Our data indicate that substantial seronegative conversion can happen in previously infected but asymptomatic individuals.

Given that most of the infected people are asymptomatic or with very mild symptoms, the short duration of the antibody response in the asymptomatic population will explain why Tokyo had the third wave. Furthermore, it can explain why the herd immunity strategy has not succeeded, and the resurgence of COVID19 is occurring every 3 to 4 months

0ur cohort was not selected from a broader, random sampling of Tokyo. Especially, our study does not include any school-age and retired elderly populations. However, due to the inclusion of a heavily commuting population in the busy area, we consider that our study population still can represent the general working population in Tokyo with high-risk exposure to the virus. Indeed, participants were well-distributed across age and gender and were sampled widely from 11 disparate locations. No documented clusters of COVID-19 among these locations are reported. Therefore, the high SPR (>40 %) is unlikely due to a cluster of COVID-19 happening in one place. More importantly, the initial SPR for this cohort started low at 5.9 % (95%CI [0,12.3%]) and jumped up beyond 40 %, mirroring the pattern of confirmed case numbers increase seen in Tokyo. These observations strongly suggest that the SPR observed in our cohort represents the viral spreading among Tokyo’s general population. However, discrete studies have to be done to determine the viral load among younger and older people. Japan took the atypical step of not instituting a mandatory lockdown during the second wave. During that time, businesses, restaurants, and transportation were kept open, and public life continued relatively unabated due to no mandatory order. Such a high seropositivity rate in Tokyo may not be entirely unexpected given its remarkably high population density, tight-spacing, the widespread use of public transportation, and no implementation of a “lockdown”.

This study is unique in utilizing a sequential serological testing approach in a moderately sized cohort of asymptomatic individuals. If the high SPR is confirmed in a more general population in Japan, the reason for remarkably low mortality related to COVID-19 in Japan should be investigated. Much like our cohort, which had no reported hospitalizations, clinical severity in Tokyo was low during the second wave. Future more extensive cohort studies are needed to confirm our results. We should evaluate what factor makes fatality low, such as lifestyle/habits, the widespread use of masks, viral mutations, and/or host factors such as immunological memory against COVID-19.

This study has some limitations. First, the cohort was a sampling of a single large company in Tokyo and not of the population, in general. Second, detailed medical histories and behavioral patterns of each employee were not obtained. Such information would have helped understand the role of cross-exposure and the factors associated with reduced fatality. Third, our antibody test is qualitative and not quantitative. Therefore, negative results indicate neither the complete loss of antibody nor of immune memory. Further studies using a quantitative antibody measurement are required.

Although the COVID-19 vaccine provides a more robust and persistent antibody response8,9, extensive coverage of the general population by vaccination is critical to eradicating COVID-19 from the community, especially if the antibody response is generally short-lived.

## Data Availability

upon the request

## Article information

### Author Contributions

All authors had full access to all the data in the study and take responsibility for the integrity of the data and the accuracy of the data analysis.

Concept and design: K Hayashida.

Acquisition of data: S Hibino, Y Hayashida

Analysis, or interpretation of data: K Hayashida, A Ahn

Drafting of the manuscript: K Hayashida, A Ahn.

Critical revision of the manuscript for important intellectual content: All authors.

Administrative, technical, or material support: S Hibino, Y Hayashida

Supervision: All authors.

